# Treatment preferences among people at risk of developing tuberculosis: a discrete choice experiment

**DOI:** 10.1101/2023.12.20.23300332

**Authors:** Wala Kamchedzera, Matthew Quaife, Wezi Msukwa-Panje, Rachael M Burke, Liana Macpherson, Moses Kumwenda, Hussein H Twabi, Matteo Quartagno, Peter MacPherson, Hanif Esmail

## Abstract

Diagnosing and treating tuberculosis (TB) early, prior to bacteriological conformation (e.g. bacteriologically-negative but radiologically-apparent TB) may contribute to more effective TB care and reduce transmission. However, optimal treatment approaches for this group are unknown. It is important to understand peoples’ preferences of treatment options for effective programmatic implementation of people-centred treatment approaches.

We designed and implemented a discrete choice experiment (DCE) to solicit treatment preferences among adults (≥18 years) with TB symptoms attending a primary health clinic in Blantyre, Malawi. Quantitative choice modelling with multinomial logit models estimated through frequentist and Bayesian approaches investigated preferences for the management of bacteriologically-negative, but radiographically-apparent TB.

128 participants were recruited (57% male, 43.8% HIV-positive, 8.6% previously treated for TB). Participants preferred any treatment option compared to no treatment (odds ratio [OR]: 0.17; 95% confidence interval [CI]: 0.07, 0.42). Treatments that reduced the relative risk of developing TB disease by 80% were preferred (OR: 2.97; 95% CI: 2.09, 4.21) compared to treatments that lead to a lower reduction in risk of 50%. However, there was no evidence for treatments that are 95% effective being preferred over those that are 80% effective. Participants strongly favoured the treatments that could completely stop transmission (OR: 7.87, 95% CI: 5.71, 10.84), and prioritised avoiding side effects (OR: 0.19, 95% CI: 0.12, 0.29). There was no evidence of an interaction between perceived TB disease risk and treatment preferences.

In summary, participants were primarily concerned with the effectiveness of TB treatments and strongly preferred treatments that removed the risk of onward transmission. Person-centred approaches of preferences for treatment should be considered when designing new treatment strategies. Understanding treatment preferences will ensure that any recommended treatment for probable early TB disease is well accepted and utilized by the public.

## Introduction

Globally, at least 10 million people fall ill with tuberculosis (TB) every year, and an estimated 1.6 million died of TB in 2022 (1). Although some progress has been made in reducing TB incidence and death, particularly in countries that have experienced HIV-driven epidemics in Africa, this has fallen well short of global targets, and has further been worsened by the disruptions to diagnosis and treatment caused by the COVID-19 pandemic (2,3). Additionally, the TB epidemic has become increasingly concentrated in marginalised groups as the overall TB prevalence has reduced (4,5). Therefore, new, person-centred, and effective approaches must be considered to diagnose and treat individuals early.

Accelerated approaches are needed to achieve the rapid declines in TB transmission required to eliminate TB. The standard approach to diagnosing disease is through bacteriological confirmation of *Mycobacterium tuberculosis (Mtb)* in a biological specimen. It is increasingly being recognised that there are potential opportunities to diagnose and intervene earlier in the disease process prior to bacteriological conformation which may then contribute to more effective TB prevention and care and interruption in TB transmission (6,7). There are several tests that have been proposed that could identify people during this earlier stage of disease some of which are established, with others currently in development. People with chest X-ray (CXR) signs suggestive of TB, but with negative sputum bacteriology, have an annualised risk of progression to bacteriologically-positive TB of 10%, with an estimated 26% progressing to sputum positivity over 3 years (8). Blood transcriptional signatures are predictive of disease progression over 0-12 months with a positive predictive value of (PPV) of 8.3-10.3% (9).

The development of diagnostics that can predict disease progression has been identified as a research priority by WHO (10). However, in recent years, there has been little attention given to the appropriate treatments for people with earlier stages of TB. Firstly, they may have more limited disease requiring less intense or shorter treatment duration than is used for bacteriologically positive disease (11). Second, a positive test predictive of disease progression does not guarantee disease progression. Analyses of historic trials providing treatment to people with CXR changes suggestive of active TB with negative bacteriology showed number needed to treat to prevent a case of bacteriologically positive TB was 2.5, which is considerably lower than that for preventive treatment of latent TB infection in a general population(12). Modern clinical trials to determine the optimal approach to treatment for probable early TB are needed. However, whilst these are being undertaken (13), it is important to better understand the treatment preferences of people in these earlier stages of TB disease to inform the design of these trials and facilitate more effective programmatic implementation of new treatment approaches.

We therefore conducted the Radio+ TB Study in Malawi to investigate the role of digital chest radiography with computer-aided detection of TB (DCXR-CAD) and a novel transcriptional marker for TB diagnosis and prognosis. Embedded in this study, we conducted a discrete choice experiment (DCE). DCEs are a widely-used tool to measure patients’ preferences for service characteristics (14–16). They have been shown to accurately predict individuals’ preferences and choices regarding healthcare services (17). In DCEs, study participants make several choices between interventions (alternatives) that have different levels of the same characteristics (attributes). It is assumed that individuals derive utility from these individual attributes, and thus their preferences can be analysed through studying their choices in DCEs(18).

Using a DCE, we aimed to investigate people’s preferences for the management of the scenario of having bacteriologically-negative sputum but with a moderate to high risk of future progression to bacteriologically positive TB.

## Materials and methods

We used a cross-sectional DCE study design to elicit people’s preferences for the management of early TB disease.

### Study setting & population

The study took place at a busy urban primary healthcare centre in Blantyre, Malawi (Bangwe Health Centre). The annualised TB case notification rate in Blantyre was 152 per 100,000 in 2020 (19). In Blantyre 15% of adults aged 18 years or older are living with HIV (20). TB diagnostic services available at Bangwe Health Centre follow standard Malawi Ministry of Health guidelines (21). Sputum Xpert MTB/Rif testing, TB treatment provision, and HIV testing and treatment were available onsite, with all clinical care provided by nurses and clinical officers, with no physicians available. At the time of the study, there were no treatment guidelines for early TB disease at the clinic. People with signs and symptoms of TB but bacteriologically-negative sputum typically do not start TB treatment, although on occasion might be referred to a tertiary hospital for further investigations or have empiric TB treatment started at the primary healthcare clinic (at the discretion of the treating nurse or clinical officer).

The present study recruited participants enrolled in the Radio+ TB diagnostic study, which included adults of at least 18 years of age, irrespective of HIV status, presenting to the study clinic with TB symptoms (cough of any duration) and residing in Blantyre. For this cross-sectional DCE survey, 128 Radio+ TB study participants were consecutively recruited after clinical assessment, which included CXR analysed with CAD4TBv6, sputum investigations, and blood testing with a TB Host Response blood test developed by Cepheid (Sunnyvale, Ca, USA), but before any results or treatment were provided. Therefore, whether or not the participant actually had bacteriologically-negative sputum and abnormal CXR was not part of the inclusion criteria.

### Design and development of the DCE tool

#### Identification of attributes

To design the DCE tool, attributes of TB management were systematically identified. An initial list of attributes was obtained from reviewing literature for the management of TB disease and treatment preference studies. We kept our search broad and did not limit the list to the management of people with a high risk of TB disease, as the literature in this area is limited and treatment guidelines for this group have not been clearly defined. The list was further informed by policy documents such as Malawi National TB and Leprosy Elimination Programme manuals and TB management guidelines.

To further inform the list of attributes and levels, we conducted qualitative interviews to ensure options were appropriate for the context in which the study would take place. The aim of these qualitative interviews was to identify important factors of the management of early TB disease. In-depth interviews were conducted with participants enrolled into the wider diagnostics study. Participants were purposively sampled to achieve a 1:1 ratio of men to women. Saturation of information was reached at 20 participants. Data were iteratively coded using Nvivo 12, and thematic analysis was employed. Study procedures and results are reported elsewhere. Table 1 illustrates the final list of attributes and levels.

**Table 1:** Final list of attributes and levels.

| Attribute | Explanation presented to the participant | Attribute Levels |
| --- | --- | --- |
| Duration of treatment | The length of time you would have to take treatment if you were at risk of developing TB disease. | - 2 months<br>- 3 months<br>- 4 months<br>- 5 months<br>- 6 months |
| Number of tablets that should be taken per dose | How many tablets you would have to take per dose | - 2 tablets per dose.<br>- 4 tablets per dose.<br>- 6 tablets per dose. |
| The reduction in the risk of being unwell with TB disease, after completing treatment | Your risk of TB disease after completing treatment will reduce from 30% to 15% (if attribute level is a 50% reduction). | - 50%<br>- 65%<br>- 80%<br>- 95% |
| Likelihood of infecting others | Whether you can still infect others after completing treatment. | - Does not stop you from passing TB on to others.<br>- Reduces the chances of passing TB on to others by half.<br>- Will completely stop you from passing TB on to others. |
| Adverse effects from the treatment | Possible harmful side effects from the treatment. | - No side effects.<br>- Minimal side effects -generally not noticeable such as brief feeling of sickness.<br>- Mild side effects present each day – may make you feel like not enjoying socializing but able to work.<br>- Moderate side effects – Bad enough to stop work and see nurse/doctor for help. |
| Frequency of follow up irrespective of treatment. | How often you would have to be checked on by health workers. | - None<br>- Once a month<br>- 3 times a month |
| Annual travel cost to access care (USD\$)* | How much you would spend travelling from your home to a healthcare facility to access care in a year. | - \$0<br>- \$2.95<br>- \$7.38 |

\*Costs in the DCE tool were expressed in Malawi Kwacha (MKW).USD\$1.00 = MKW812.51 as of

#### Experimental design and construction of DCE choice sets

In constructing the choice sets for the DCE, using a full factorial design would be the most balanced and orthogonal as it contains all possible combinations of attributes and levels (23). However, with a total of 7 attributes and 25 levels this would result in 1024 (4^3^ X 2^4^ X 1^5^) choice sets, which was impractically large and would result in respondent fatigue (23). Therefore, to make the task manageable, we used a fractional factorial design (23,24), whereby participants were exposed to a random selection of 10 choice sets from the full factorial design.

Using the initial choice set we conducted two iterations of cognitive interviews among five participants. We aimed to investigate whether the formulated DCE was comprehensive for the target population. This involved discussions around the presentation and format of the DCE, including the visual aids and wording used. The results of this were then used to iterate and improve upon the DCE tool.

Thereafter, we conducted a pilot study among a further five participants to ensure that the most meaningful and efficient design was used for the final study. To ensure the most efficient DCE design was formed, we used NGENE software to formulate different designs that minimised D-error(23). The D-efficiency score refers to the precision with which effects of attributes are estimated in the model (24). Formulation of D-efficient choice sets depends on the planned model specification and the assumed parameter values (23). We therefore used the parameters estimated with the pilot data to act as the assumed parameter values to inform the formulation and selection of the most D-efficient design. We also used the piloting process to identify any areas of the DCE’s format and the respective visual aids that needed further improvement.

The final DCE included 10 choice tasks. We also included an opt-out option of “neither” to ensure the choices made are realistic in allowing patients not to choose treatment. Each choice task included a scenario, where the participant had negative sputum bacteriology and a CXR result suggestive of TB and underwent a further (unspecified) predictive tests with the result providing a hypothetical risk of disease progression. Participants were therefore asked to make their choices between treatments based on each scenario (e.g.: “if you had a positive test result which means your risk of developing TB disease over the next 12 months is xx%, which treatment would you prefer?”). The risk level of disease progression varied with each choice set (50%, 30%, and 10%). This was done to understand whether the known level of disease risk due to a predictive test could influence treatment preferences. We developed visual aids from the cognitive interviews, which were used to explain the meaning behind the “risk of TB disease progression” and the attributes. To check consistency in choices we randomly picked one choice task to be repeated at the end of the 10 tasks. We also asked the participants to give a reason as to why they have picked a particular treatment option. This was done to ensure choices made were not random or due to decision fatigue (25).

### Data analysis

Data were analysed using STATA v14 (College Station, Texas, US) and R version 4.2.2 (R Foundation for Statistical Computing, Vienna, Austria). Descriptive statistics were used to summarise the data and to investigate the distribution of key participant characteristics within the study population.

We employed quantitative choice modelling by using a multinomial logit model to analyse the choices made by the participants. In this model the dependant variable was the choice made by participants between treatments, and independent variables were the different attributes and their respective levels. In such models, it is assumed that individuals derive their utility or satisfaction from the choices made between treatments (18). Therefore, the model-derived coefficients for the different attribute levels provides the marginal effect of each attribute level on the choice of treatment for individuals at risk of TB disease progression, holding all other attributes constant. Coefficients from the multinomial logit model were converted to odds ratios to show the relative effect of each attribute level on the choice of treatment as compared to the attribute’s base level.

We further evaluated the interaction between the risk of TB disease progression, taken from the hypothetical scenarios, and the different attribute levels to investigate whether the effect of the attributes on preferences varies by the risk of disease progression. Due to there being an insufficient sample size to observe variations according to disease risk, we employed Bayesian methods to construct a multinomial logit model to estimate possible interaction effects. Priors were weakly regularising. We summarised posterior draws to estimate the percentage point difference and uncertainty in the probability of selecting a treatment option due to an attribute level.

### Ethical approval

Ethical approval was received from the University of Malawi College of Medicine Research Ethics Committee (P.02/21/3271) and University College London Research Ethics Committee (ID Number: 19219/001). All participants provided written informed consent. If illiterate, participants were asked to provide a witnessed thumbprint.

## Results

Participants were recruited from the 14^th^ of September 2021 to the 21^st^ of February 2022. Table 2 shows the study participants’ characteristics. Of the 128 participants, 57% were male. Most participants were aged between 35 and 44 years (30.5%). Additionally, most participants (84.4%) reported primary education as their highest level of education. 40.6% of participants were self-employed and 23.4% reported having formal employment. The median duration of cough experienced by participants before presenting to the clinic was 21 days (IQR: 14-30). After cough, chest pain was the most common symptom experienced by participants (60.9%). Approximately 8.6% of participants had been previously treated for TB. Just under half of the participants reported being HIV positive (43.8%). During the main study, 10.2% of the participants initiated TB treatment over 6-months of follow-up.

**Table 2:** Participant Characteristics N=128.

| Characteristic |  | N (%) |
| --- | --- | --- |
| Sex | Male | 73 (57.0) |
|  | Female | 55 (43.0) |
| Age in years | 18-24 | 24 (18.8) |
|  | 25-34 | 34 (26.6) |
|  | 35-44 | 39 (30.5) |
|  | ≥ 45 | 31 (24.2) |
| Marital status | Married/cohabiting | 86 (67.2) |
|  | Single/ widowed | 42 (32.8) |
| Highest education level obtained | None | 6 (4.7) |
|  | Primary | 108 (84.4) |
|  | Secondary (MSCE) | 10 (7.8) |
|  | Higher education | 4 (3.1) |
| Literacy | Can read | 116 (90.6) |
|  | Cannot read | 12 (9.4) |
| Employment | Employed | 30 (23.4) |
|  | Self-employed | 52 (40.6) |
|  | Student/other | 3 (2.3) |
|  | Unemployed | 43 (33.6) |
| Median duration of cough in days (IQR) |  | 21 (14-30) |
| Experienced other TB symptoms | Chest pain | 78 (60.9) |
|  | Dyspnoea | 57 (44.5) |
|  | Feeling weak | 51 (39.8) |
|  | Fever | 54 (42.2) |
|  | Sweating | 52 (40.6) |
|  | Weight loss | 74 (57.8) |
| Previously treated for TB | Yes | 11 (8.6) |
|  | No | 117 (91.4) |
| Self-reported HIV status | Positive | 56 (43.8) |
|  | Negative | 65 (50.8) |
|  | Missing | 7 (5.5) |

MSCE: Malawi School Certificate of Education IQR: Interquartile range. TB: tuberculosis

Table 3 shows the results of the initial multinomial logit model. The presented odds ratios show the odds of choosing a treatment due to an attribute level compared to the attribute’s base level.

**Table 3:** Treatment preferences N=128.

| Attribute | Level | Odds Ratio (95%CI) | *P-value |
| --- | --- | --- | --- |
| Number of tablets per dose | 2 | 1 |  |
|  | 4 | 0.83 (0.63, 1.08) | 0.17 |
|  | 6 | 0.60 (0.40, 0.92) | 0.02 |
| Duration of treatment (months) |  | 1.41 (1.28, 1.56) | <0.001 |
| Reduction in risk of developing disease after completing treatment | 50% | 1 |  |
|  | 65% | 0.91 (0.67, 1.24) | 0.57 |
|  | 80% | 2.97 (2.09, 4.21) | <0.001 |
|  | 95% | 1.95 (1.33, 2.85) | 0.001 |
| Likelihood of infecting others with TB disease | Does not stop transmission | 1 |  |
|  | reduces the chances of transmission by half | 8.4 (6.12, 11.54) | <0.001 |
|  | Completely stops transmission | 7.87 (5.71, 10.84) | <0.001 |
| Adverse effects | None | 1 |  |
|  | Minimal | 0.35 (0.25, 0.50) | <0.001 |
|  | Mild | 0.22 (0.16, 0.32) | <0.001 |
|  | Moderate | 0.19 (0.12, 0.29) | <0.001 |
| Follow up | No follow up | 1 |  |
|  | Once a month | 2.57 (1.99, 3.33) | <0.001 |
|  | 3 times a month | 1.31 (1.01, 1.72) | 0.05 |
| Annual travel cost | No cost | 1 |  |
| | USD\$2.95 | 2.02 (1.04, 3.93) | 0.037 |
| | USD\$7.38 | 3.49 (2.15, 5.67) | <0.001 |
| Opting out of treatment |  | 0.17 (0.07, 0.42) | <0.001 |

CI: Confidence Interval
USD\$1.00 = MWK812.51 as of 14/09/22(22)

There was strong evidence that people valued any treatment option over no treatment (p<0.001). Compared to treatments that did not stop transmission, treatments that reduced the chances of transmission by half had 8.4 (95% confidence interval [CI] 6.12, 11.54) times greater odds of being chosen, and those that completely stopped transmission had 7.87 (95% 5.71, 10.84) times greater odds of being chosen. Participants strongly preferred treatments that reduced their risk of developing TB disease by 80% (odds ratio [OR]: 2.97; 95% CI: 2.09, 4.21) and 95% (OR: 1.95; 95% CI: 1.33, 2.85) compared to treatments that lead to a reduction in risk by 50%.

There was strong evidence for participants being opposed to treatments with moderate (OR: 0.19, 95% CI: 0.12, 0.29), mild (OR: 0.22, 95% CI: 0.16, 0.32) and minimal side effects (OR: 0.35, 95% CI: 0.25, 0.50) compared to treatments with no side effects. There was evidence that participants did not prefer treatments that had 6 tablets per dose as compared to those with 2 tablets per dose (OR: 0.60; 95% CI: 0.40, 0.92). Compared to treatments that did not require any follow-up visits, participants preferred treatments with follow-up visits taking place once a month (OR: 2.57; 95% CI: 1.99, 3.33).

There were some counter-intuitive results. With an increase in the number of months required to complete treatment, the odds of participants preferring the treatment also increased (OR: 1.41; 95% CI 1.28, 1.56). Additionally, participants preferred treatments with a higher annual travel cost as compared to those that incurred no cost (OR: 3.49; 95% CI: 2.15, 5.67).

To check the consistency of participants’ choices, task 5 was repeated at the end of all the choice sets in the DCE. Of the 128 participants 80.5% chose the same treatment option.

To explore whether quantification of the risk of TB affected the importance of different attributes, the interaction effect between the risk of TB disease progression and the different treatment attributes were modelled using a Bayesian multinomial logit model. The results of this are illustrated in Figure 1. Generally, the results showed that, regardless of the level of disease risk, participants were more likely to opt for treatment rather than opt out of treatment. There was, however, no significant evidence for a difference in preferences according to the disease risk level for all treatment attributes.

**Fig 1:**
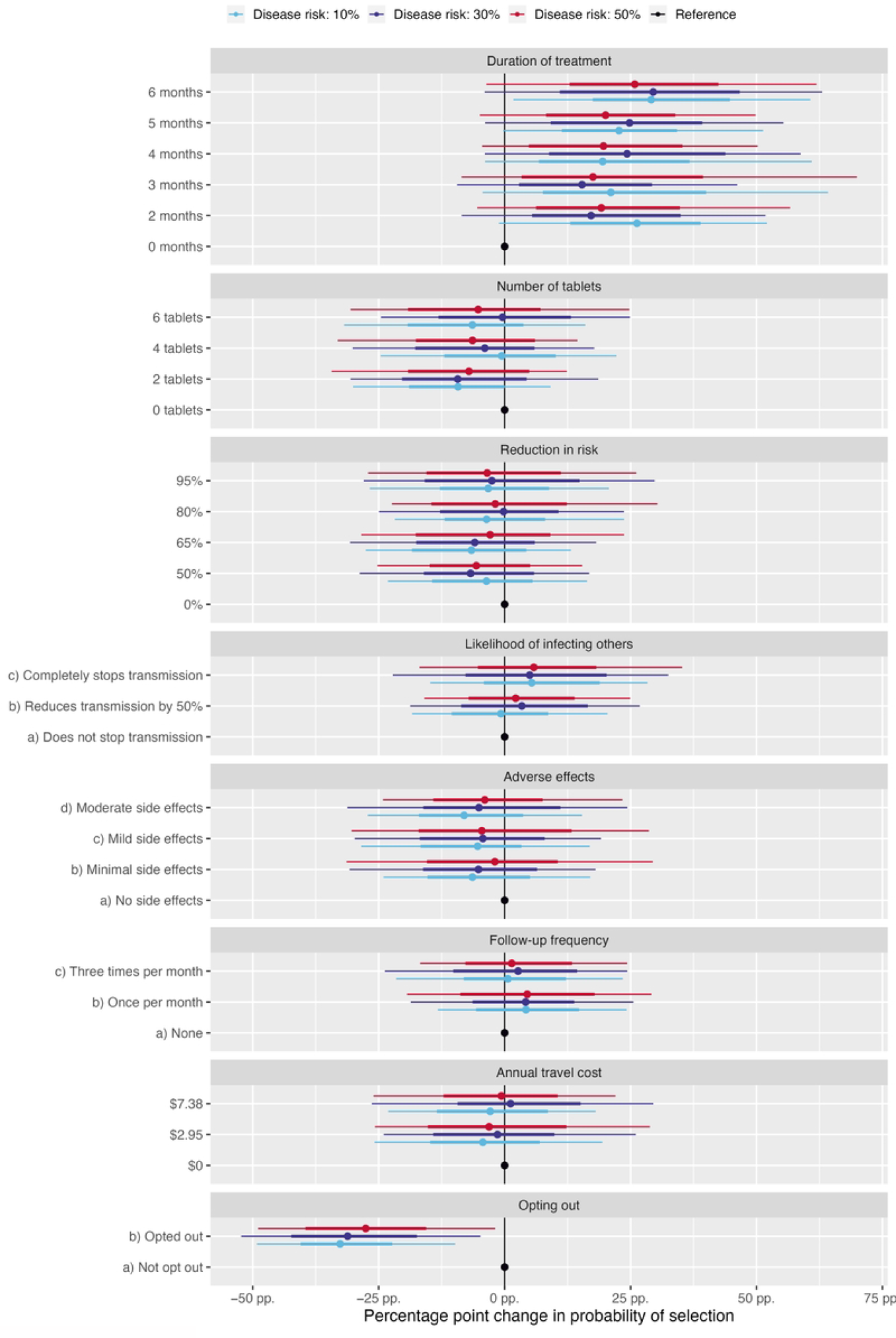
Interaction effect between risk of TB disease progression and treatment attributes N=128 USD$1.00 = MWK812.51 as of 14/09/22(22)

## Discussion

We set out to investigate people’s preferences for the treatment in the scenario of not having disease bacteriologically confirmed but being presented with a high risk of future disease progression. Our main findings were that, among those at risk of progression to clinical TB, there is a strong preference to take treatment than to not take treatment. This finding was regardless of the level of perceived TB disease risk. Furthermore, strong preferences were shown for treatments that were highly effective, such as treatments preventing transmission and those that would likely reduce the risk of disease progression. The main implications of these findings are that to ensure treatment uptake, new treatment strategies should be highly effective both in treating disease and in reducing the risk of transmission. Additionally, for novel treatments that do provide these benefits, it is important to raise and ensure awareness of these benefits among those seeking healthcare to ensure treatment uptake and adherence.

In our study, participants were shown to be primarily concerned with cure. This is demonstrated by the preference of being on any treatment option as opposed to none; and there being a strong preference for treatment options that reduced their risk of TB disease by at least 80% as compared to treatments offering a 50% reduction. This is further illustrated with there being no interaction effect between the risk of disease progression and attribute levels. This is in line with previous studies on preferences for both preventative therapy and treatment for TB as people have been shown to prioritise being completely cleared from infection or disease, and expressed that their motivation for adhering to treatment is the desire to be healthy (26–28).

Preferences were strongly dominated by treatment options that reduced the likelihood of *Mtb* transmission. Previous studies employing qualitative interviews and DCEs have shown that a motivating factor for many individuals to initiate preventative TB treatment was to protect close contacts such as family members, especially children (27). Another study looking at public preferences for interventions that prevent infectious diseases demonstrated that most participants preferred control measures such as isolation and quarantine in order to prevent the spread of infection to others (29). Altogether, this suggests that for our study population, protecting others from infection was a main priority.

Concerning the features of treatment, preference for a treatment increased with an increase in the duration of treatment. This is against our expectations as previous studies have demonstrated that people prefer treatments of shorter durations and the reasons for this is to minimise the risk of side effects to ensure treatment adherence (27,30). With our DCE study population, it is possible that participants preferred longer durations as opposed to shorter durations because of a lack of confidence in shorter treatments being effective. This may have been influenced by the current TB treatment regimen in Malawi being six months and the participants’ strong preference for treatments that are highly effective by reducing their risk of disease progression. Although it is possible that participants were unable to understand the attribute within the DCE or the design of the DCE was not able to effectively measure the participants’ preferences, we conducted cognitive interviews before the study to minimise this risk. Further qualitative research is needed to validate and improve our understanding of this result.

Our results showed that the experience of taking medication was of high priority. There was preference for treatments with a lower pill burden and no to minimal side effects. A previous study also found that the pill burden of TB treatment had a negative impact on the patient’s experience and therefore treatment adherence (28). Similarly, previous studies have shown that people viewed side effects as a barrier to treatment adherence (26,27,30). Side effects arising from TB treatment may hinder an individual’s quality of life and ability to take part in income generating activities whilst being on treatment. This is crucial for households in low-income countries such as Malawi.

Participants also preferred to have routine follow-up whilst on treatment. However, the optimal frequency of follow-up was once a month. We can speculate that this finding may reflect the awareness of barriers to accessing healthcare in low-income countries, such as distance and poverty (31–33). And once they are at the health facility, individuals often face long waiting times and high costs (33–35).

Guo *et al* (2011) showed that participants that were employed preferred less frequent clinic visits whilst taking TB preventive therapy (27). This may be especially true with our study population since the majority are either employed or self-employed. Overall, this suggests that, most people desire routine follow-up; however this should not come at the expense of forgoing work related or productive activities.

The odds of picking a treatment increased with cost. This was in contrast to our prior expectations and previous literature (28). It has been shown that high costs act as a barrier to accessing care, and individuals with TB often fall victim to catastrophic expenditure (32,33,36). However, participants preferring to be on any treatment as opposed to no treatment is likely to have driven these results. Opting out of treatment would entail not paying anything as treatment is not being accessed. Therefore, the choice of choosing any treatment option over no treatment would have implied that individuals would rather bear a small cost if it meant being on treatment and getting cured.

It is important to note that the participants were presented with all attributes and levels of each treatment option therefore allowing them to weigh the costs and benefits of each treatment. In reality, this is not the case, as not all benefits are made known to individuals in advance of seeking treatment. Therefore, the costs faced can still act as a barrier to seeking care despite the treatment being highly effective.

Our results show that 80.5% of the participants selected the same treatment option during the repeated choice tasks. This shows consistency in choices and is broadly in line with other DCEs in the literature, therefore demonstrating internal validity of the results (37).

## Study limitations

One limitation of this study is that it was not sufficiently powered to explore heterogeneity in people’s preferences for early TB treatment across key demographic characteristics. This type of analysis is important because it acknowledges the fact that the population is not completely homogenous and therefore preferences can be influenced by other factors. Identifying these influencing factors can give us a greater understanding of why different treatments and interventions are accepted at varying degrees in different populations.

This study only recruited individuals with TB symptoms presenting to the clinic. However, it is becoming apparent that a significant proportion of individuals with early TB disease can be asymptomatic. As these participants are not experiencing any symptoms, they may not anticipate being put on treatment for an extended period. Therefore, their preferences for treatment may be different from those that are symptomatic. Future studies should explore the preferences of this group as insights from such studies can give further information on how patient-centred approaches for early TB management can be designed.

Although we asked the participants why they preferred certain treatment options to ensure choices made were not random or due to fatigue, we did not formally record the decision-making process of our study’s participants when choosing between treatment options. This data would have been valuable because it would provide further evidence as to why certain treatment attributes were preferred over others. Therefore, we were only able to speculate as to why certain choices were made when discussing the results of this study. Future DCEs should consider this practice to add depth to the results obtained.

## Conclusions

Our study demonstrates that for the treatment of individuals with a risk of TB disease progression, there is a primary concern with the effectiveness of treatment. Additionally, people will strongly consider taking treatment if it means reducing or removing the risk of transmission. Therefore, new treatment strategies should ensure that the treatment is highly effective in treating TB disease. Furthermore, our results also have implications on how public health messages around new treatment strategies should be presented and packaged to the public. For instance, if people are aware of the reduction in the risk of TB transmission whilst being on treatment they are more likely to desire early TB treatment and therefore achieve individual and community benefits. Concerning treatment characteristics, the persons experience should be considered when designing new treatment strategies as people preferred no to minimal side effects, and fewer tablets per dose. Altogether, this will ensure the treatment is well accepted and utilized by the public.

## Data Availability

The data dictionary and full dataset has been uploaded to the UCL research repository (private link https://figshare.com/s/2c63e73d69d6e7f58916). Embargo date 1/3/2024 which will be aligned with publication date

## Supporting information

S1 Appendix. Discrete choice experiment questionnaire (English)

S2 Appendix. Discrete choice experiment questionnaire (Chichewa)

S3 Appendix. Discrete choice experiment visual aid.

